# Detection of infectious SARS-CoV-2 in frozen aerosol samples collected from hospital rooms of patients with COVID-19

**DOI:** 10.1101/2022.11.14.22282295

**Authors:** Audray Fortin, Marc Veillette, Adriana Larrotta, Yves Longtin, Caroline Duchaine, Nathalie Grandvaux

## Abstract

We isolated infectious SARS-CoV-2 from aerosol samples collected from hospital rooms of COVID19 patients. Isolated virus successfully replicated in cell cultures 14 months after collection, opening up prospects for retrospective analyses of samples stored during the previous waves of COVID-19.

## Introduction

At the stage of the COVID-19 pandemic where vaccines have greatly contributed to limiting the severity of the disease and the pressure on health systems, surveillance methods allowing the assessment of community levels of SARS-CoV-2 remain essential to inform decisions about strategies to prevent COVID-19 transmission. Understanding virus emission rate and subsequent transmission of SARS-CoV-2 through the air via particles of different sizes, generally called droplets and aerosols, implies that we have well-defined methods to monitor indoor air [1] [2]. This is essential to better understand viral resistance to environmental stress, inform on the risks of acquisition in the community and occupational environments and appropriately evaluate virus mitigation methods in indoor settings. Several studies have reported the presence of SARS-CoV-2 RNA in air samples in various settings such as long-term care, acute and intensive care, cars and homes [2-5]. Unlike molecular methods that allow the quantification of viral genome in air samples, measuring infectious viral particles that may contribute to SARS-CoV-2 transmission is not trivial and has been a technical challenge knowing that the samplers, the environmental context, the time of sampling and the need to store the samples before analysis can strongly influence the quantity of viral particles collected as well as their infectivity [2]. Demonstrations of the presence of infectious viruses capable of replicating in cells have been rarer [3, 6, 7]. In this study, we evaluated the capacity to isolate replicating SARS-CoV-2 from environmental aerosol samples from a hospital setting after air sample freezing and long-term storage.

## Methods

### Bioaerosols collection and processing

Two types of sampling devices were used. First, 37mm cassettes with 0.8µm polycarbonate filters (SKC, Eighty Four) were located at the head of the beds at 1.5-2.m from the patient’s head and oriented face down. Second, a Series 110A Liquid Spot Sampler (Aerosol Devices), which consists of a growth tube of water condensation followed by a single jet collector impactor in a liquid, was located at 2-3m from the patient’s bed (**Supplemental Figure 1 and methods**). Samples in Viral Transport Media (VTM, Redoxica) were stored at -80°C.

### SARS-CoV-2 replication in cell culture

All experiments related to SARS-CoV-2 culture were conducted in a certified containment level-3 (CL3) facility, using standard operating procedures approved by the Biosafety committee at CRCHUM, Montreal, Canada. Previously frozen air samples in VTM were used to inoculate VERO E6 cells for 2 successive rounds of infection as detailed in **Figure 1E**. Live or β-propiolactone (BPL) inactivated SARS-CoV-2/SB2 isolate [8] were used as control. Cells and supernatants were collected to assess parameters of virus replication. Cytopathic effects (CPE) were assessed through brightfield microscopy using an EVOS FL Auto 2 microscope (Life technologies). Virus titer in supernatants was assessed by median tissue culture infectious dose (TCID50). SARS-CoV-2 protein detection in cells was performed by analysis of whole cell extracts by immunoblot using anti-Spike and anti-Nucleocapsid antibodies (**Supplemental methods**).

**Figure 1.**
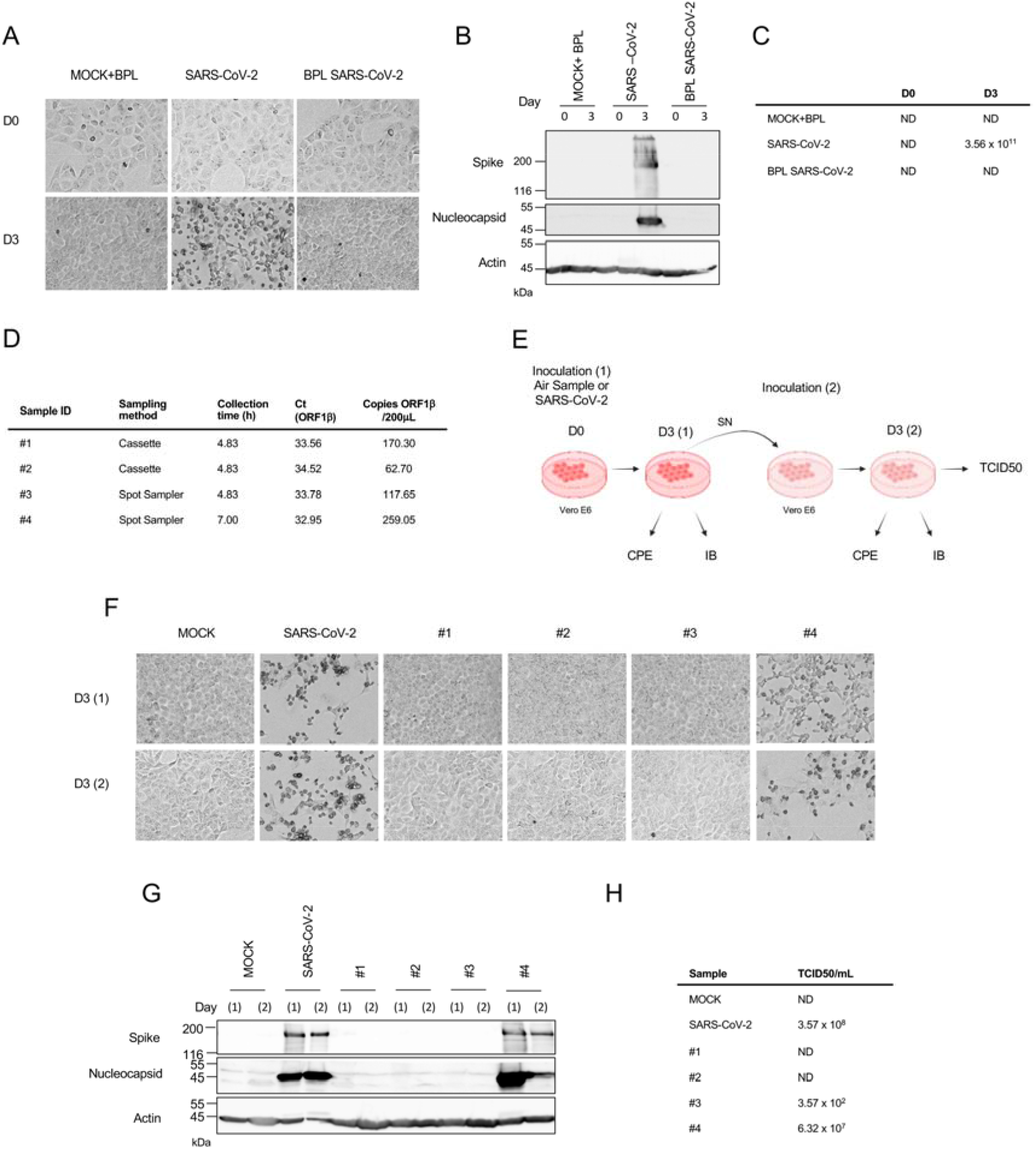
**(a-c)** Test experiment to evaluate the cytopathic effects (CPE), expression of Spike and Nucleocapsid and *de novo* virions production occurring in VERO E6 cells infected with 150pfu of replicating and Beta propiolactone (BPL)-inactivated SARS CoV-2. Parameters were evaluated at Day 0 (D0) and day 3 (D3) of infection. **a)** CPE was detected by brightfield microscopy. **b)** Cellular expression of SARS-CoV-2 spike and nucleocapsid proteins was detected by immunoblot. **c)** Production of *de novo* virions in the supernatant of infected cells was quantified by the median tissue culture infectious dose (TCID50) calculation method. **d-h**) Analysis of air samples collected in COVID-19 patients hospital rooms. **d**) Quantification of SARS-CoV-2 ORF1b RNA by RT-qPCR. **e**) Experimental scheme used to culture SARS-CoV-2 from air samples. Air samples were used as inoculum on VERO E6 cells. After 3 days of infection, the supernatant was collected and used to inoculate fresh VERO E6 cells. Control infection was performed with 150 pfu SARS-CoV2. CPE (**f**), cell expression of spike and nucleocapsid proteins (**g**) and production of *de novo* virions (**h**) were monitored. ND: not detectable.

### RT-qPCR

RNA from air samples or cell supernatants were subjected to RT-qPCR for amplification of SARS-CoV-2 ORF1b or N as detailed in **Supplemental methods**. The estimation of the quantity of SARS-CoV-2 genome per cubic meter of air was extrapolated based on the ORF1b quantification data.

### Patient characteristics

The characteristics of patients occupying the rooms with detectable airborne infectious virus were extracted by reviewing their medical chart. The study was approved by the ethics committee of the Institut Universitaire de Cardiologie et de Pneumologie de Québec (IUCPQ) under the project identifier MEO-21-2021-3475. A waiver of individual written informed consent was approved for the air samplings and the retrospective extraction of patient information from medical records.

## Results

Air samples were collected in acute care hospital rooms located in a unit dedicated to the care of patients with COVID-19 between October, 27 2020 and November, 6 2020 in the Quebec province, Canada when mostly the Alpha variant of concern was in circulation and before vaccines were available. A total of 30 samples were collected in 8 different rooms occupied by patients with acute COVID-19 using cassettes or a Spot Sampler devices. The sampling duration ranged from 4.75h to 7.0h (mean=6.18h). A total of 9/22 (40.9%) cassettes and 2/8 (25%) Spot Sampler samples were found positive for SARS-CoV-2 RNA as determined by RT-qPCR, with airborne concentrations ranging from 129 to 2056 genomes equivalent per cubic meter of air (mean=644).

First, to establish that our cell culture design differentiated between replicating and non-replicating virus, VERO E6 cells were inoculated with 150pfu of replicating (chosen based on the mean copy number of the air samples analyzed in cell culture) or BPL-inactivated SARS-CoV-2/SB2 isolate. Three days after infection, SARS-CoV-2 replicating virus induced significant signs of CPE (**Figure 1A**), analysis of whole cell extracts by immunoblot allowed detection of spike (S) and nucleocapsid (N) proteins (**Figure 1B**), and quantification of the supernatant revealed high levels of *de novo* virions (**Figure 1C**). Importantly, none of these parameters were positive when monitored 2 hours after inoculation of replication-competent virus, or when using BPL-inactivated SARS-CoV-2 as inoculum. These results confirmed that only actively replicating virus causes a CPE, exhibit cellular expression of S and N proteins, and lead to detectable *de novo* virions production.

Four air samples (two positive Spot Sampler samples and two corresponding positive cassettes) collected from the same patient room among the highest RNA concentrations detected were selected to test for the presence of replicating virus in cell culture. As per RT-qPCR quantification, the number of SARS-CoV-2 ORF1b copies in the aerosol samples chosen to be tested ranged from 62.70 to 259.05 in 200μL (mean = 152.43/200μL, **Figure 1D**). Patient characteristics (**Supplemental Table 1**), that may influence aerosolization of SARS-CoV-2 were acute COVID-19 pneumonia, dyspnea that required intermittent administration of oxygen by nasal canula and severe cough that required the administration of oral codeine.

The four air samples had been previously stored frozen at -80°C and were analyzed for the presence of replicating virus in cell culture in late February 2022, i.e. 14 months after collection. For this purpose, 200μl of air samples was used as inoculum to infect VERO E6 cells. Considering that air samples presented low SARS-CoV-2 particle numbers according to the RT-qPCR detection of ORF1b (**Figure 1D**), a second cycle of infection of fresh VERO E6 cells was carried out using the supernatant of the infected cells at day 3 to maximize the amplification of the virus and therefore the ability to detect low levels of viable particles (**Figure 1E**). In addition, this strategy makes it possible to confirm that SARS-CoV-2 in the inoculum has the capacity to carry out a complete replication cycle in the cells leading to the *de novo* production of infectious virions. Samples #4 collected using the Spot Sampler induced detectable CPE at day 3 after the first and second inoculations, with destruction of the monolayer inferior to that observed after infection with 150pfu of SARS-CoV-2/SB2 isolate (**Figure 1F**). Cellular spike and nucleocapsid were detected in WCE from cells inoculated with sample #4 at day 3 of the first and second infections, indicating the presence of infectious SARS-CoV-2 (**Figure 1G**). The titer of virions in the supernatant produced by sample #4 after 2 cycles of infection was of 6.32 × 10^7^ TCID50/mL, which is 5.67 times lower than the titer observed after infection with 150pfu of SARS-CoV-2/SB2 isolate (**Figure 1H**).

TCID50 analysis also allowed the detection of virions (3.57 × 10^2^ TCID50/mL) in the supernatant of cells infected with sample #3 after 2 rounds of infection, while neither CPE nor detection of spike and nucleocapsid was achieved at day 3 of the first or second infections. Neither of the two samples collected with the cassette produced detectable CPE, viral proteins expression or *de novo* virions.

## Discussion

While the detection of SARS-CoV-2 RNA in indoor air samples collected in various settings has been widely reported, only a few studies have successfully documented the presence of infectious virus [2]. Here, we successfully detected replicating SARS-CoV-2 in one out of four air samples collected in COVID-19 patients hospital rooms, 14 months after sample collection.

Our study adds to previous reports showing replicating SARS-CoV-2 virions in aerosols from COVID-19 patients’ hospital rooms [6, 7]. In this study, the infectivity of the virus present in air samples was tested in two successive rounds of cell culture inoculation using virus-induced CPE detection, immunoblotting of viral proteins and titration of infectious virions as read-outs. We used immunoblotting against the SARS-CoV-2 S and N proteins to confirm the presence of SARS-CoV-2 in cell culture experiments. Our samples were collected at time of Alpha variant of concern circulation. With the development of antibodies recognizing SARS-CoV-2 proteins of different variants of concern, this technique should be applicable with different circulating variants to demonstrate viral protein expression. RT-qPCR of the nucleocapsid RNA was also performed on the supernatant at day 3 of the second inoculation (**Supplemental Table 1**), thus further confirming the presence of SARS-CoV-2 in sample #4 that was positive in the immunoblot assay. No SARS-CoV-2 N RNA was detected in sample #3, while a 3.57 × 10^2^ TCID50/mL titer was quantified in the supernatant at day 3 of the second inoculation. Neither CPE nor viral protein immunodetection were observed with this sample, making it impossible to conclude on the presence of infectious SARS-CoV-2 in this sample.

The accumulation of data on the transport of infectious particles of SARS-CoV-2 in the air has in part been hampered by the difficulty of implementing the cell culture-based test due to the need for access to a level 3 containment laboratory suitable for SARS-CoV-2. Such studies often require air samples to be frozen and stored for varying lengths of time, which can alter virus viability after the collection step. In the present study, we demonstrate the recovery of replicative virus particles in air samples after freezing at -80°C in the VTM and storage for 14 months. It is noteworthy that during the storage period, samples also underwent two periods of transport on dry ice. Our results are of the utmost importance because they make it possible to consider the retrospective evaluation of samples taken during the different waves since 2020 for the presence of infectious SARS-CoV-2.

Out of the four samples tested, the only one from which we successfully retrieved virus capable of replicating in cell culture was collected using the Spot Sampler. This result adds to the body of evidence that demonstrates that CGT samplers allow the collection of airborne SARS-CoV-2 and better preserve virions infectivity [3, 6].

## Data Availability

All data produced in the present work are contained in the manuscript

## Acknowledgements

The authors thank Dr. Samira Mubareka (Sunnybrook Research Institute, Toronto, Canada) for the SARS-CoV-2/SB2 isolate used as a control in this study and Dr. Arinjay Banerjee (Vaccine and Infectious Disease Organization, University of Saskatchewan, Saskatoon, Canada) for advice for SARS-CoV-2 culture. Figure 1E has been created with BioRender.com

