## Supplemental information for "Detection of infectious SARS-CoV-2 in frozen aerosol samples collected from hospital rooms of patients with COVID-19"

**Supplementary methods**

***Bioaerosols collection and processing***

Samples were collected at a flow rate of 10L/min when the polycarbonate filters (SKC, Eighty Four) and 1.5L/min when using the Series 110A Liquid Spot Sampler (Aerosol Devices). Two ml of Viral Transport Media (VTM, Redoxica) were used to elute cassettes. Spot Sampler samples (100μl VTM) were completed to 800 μl using VTM. Aliquots were prepared on ice and stored at -80°C.

***RT-qPCR***

RNA extracts performed on frozen air sample were prepared using MagMAX™ Viral RNA Isolation Kit (Applied Biosystems, Vilnius, Lithuania), according to the manufacturer's instructions. RNA was eluted with 50 μL of elution buffer and stored at −80°C until quantification. RNA extractions performed on cell supernatant (140 μl) were performed using the QIAamp Viral RNA Mini Kit (Qiagen, Cat#52904). RNA was eluted in a final volume of 80μl. RT-qPCR reactions were performed as described in [1]. Briefly, the detection was performed in duplicates for both targets ORF1b and N. No template and extraction controls were tested for each batch. Positive control plasmids were included with each batch to allow detection and quantification: 2019-nCoV_N positive control plasmid from IDT and a custom plasmid with the ORF1b insert. Quantification was achieved based on the ORF1b plasmid standard curve with a limit of detection of 1 plasmid/qPCR reaction. The estimation of the quantity of SARS-CoV-2 genome per cubic meter of air was extrapolated based on the ORF1b quantification data.

***Cell Culture***

The VERO E6 cell line (ATCC, cat#CRL-1586), isolated from African green monkey kidney, was cultured in DMEM medium (GIBCO, cat#11995-065) supplemented with 1% L-glutamine (GIBCO, cat#25030-081), 10% Fetalclone III (FCl-III, Hyclone, cat#SH3010903) and the following antibiotic-antimycotic mixture: 2.5μg/mL vancomycin, 4μg/mL trimethoprim, 4μg/mL ceftazidime, 1μg/mL amphotericin B, 24.8μgmL fluconazole and 1/100 penicillin/streptomycin (Gibco, cat#15140-122). Mycoplasma contamination was regularly monitored using the MycoAlert Mycoplasma Detection Kit (Lonza). All cell culture was done in a 5% CO2 incubator at 37°C.

***SARS-CoV-2 virus culture and infection***

All protocols related to SARS-CoV-2 virus culture were performed in the Biosafety Level III laboratory at Centre de Recherche du CHUM. The SARS-CoV-2/SB2 isolate [2] was obtained from Dr. Samira Mubareka, (Sunnybrook Research Institute, Toronto, Canada) and propagated in VERO E6 cells at a MOI of 0.02. At 96h post-infection, the supernatant was harvested and clarified to remove cell debris by centrifugation at 4000g for 15min. For inactivation, the clarified supernatant was treated with 0.05% (v/v) β-propiolactone (BPL, Sigma, cat#P5648) for 16h at 4°C. Residual BPL was hydrolyzed during 2h at 37°C. Inactivation was confirmed by lack of replication in VERO E6 cells. Tests were performed on SARS-CoV-2 virus stocks to exclude mycoplasma contamination (Invivogen, cat#REP-MYS-10).

For infection experiments, VERO E6 cells were seeded at 3.5 x 10^5^ cells/well in a 12-well plate (Corning, cat#3513), to obtain 90% confluency the next day. Twenty-four hours after seeding, cells were inoculated with air samples in VTM or with 150pfu of replicating or BPL inactivated SARS-CoV-2/SB2 and left to incubate for 2h with intermittent shaking (every 15 min) for the first hour. After 2 h, the inoculum was removed, and cells were thoroughly washed three times with 2mL media containing 2% FCl-III before addition of 1mL of fresh media containing 2% FCl-III. The infection was pursued for 3 days. For conditions using air samples as inoculum, the supernatant was collected and used as inoculum to perform a 2^nd^ viral infection on fresh VERO E6 cells as described above. Infection was pursued for 3 days. Visualization of cytopathic effects (CPE) during the infection was performed through brightfield microscopy using an EVOS FL Auto 2 miPE microscope (Life Technologies). Supernatants were collected and subsequently used to evaluate viral particle titer via the median tissue culture infectious dose (TCID50). Cells were collected to assess the presence of SARS-CoV-2 proteins by immunoblot.

***TCID50 titration of SARS-CoV-2***

VERO E6 cells were seeded at 6 x 10^4^ cells/well in a 96-well plate (Sarstedt, cat#83.3429), to obtain 95-100% confluency the next day. Serial dilutions of collected supernatants were performed (10^-1^ to 10^-10^) in culture medium containing 10% FCl-III. Fifty μL from each of these dilutions along with the undiluted sample, as well as a mock control consisting of 10% FCl-III media only, were inoculated on the VERO E6 cells. Cells were left to incubate for 1h with intermittent shaking of the plate for the first hour. After 1h, the inoculum was removed and replaced with fresh media containing 2% FCl-III. After a 5-day incubation in a 5% CO_2_ incubator at 37°C, virus titer calculation was performed based on CPE evaluation in each well combined with the Spearman & Kärber algorithm [3, 4].

***Immunoblot analysis.***

Cells were harvested in cold DPBS (Gibco), placed into pre-chilled 1.5mL tubes and centrifuged at 16200g for 30sec. Cell pellets were then resuspended in 11.25µL RIPA lysis buffer composed of 50mM Tris pH 7.5, 150mM NaCl, 2mM EDTA, 0.1% SDS, 1% Triton and 1% Sodium Deoxycolate to which 10μg/mL leupeptin, 10μg/mL aprotinin, 10μg/mL pepstatin, 30mM NaF, 1mM activated Na_3_VO_4_, 1mM p-nitrophenyl phosphate and 10mM β-glycerophosphate were added. Samples were left to incubate 20min on ice and then centrifuged at 16200g for 20min. Supernatants were collected in new pre-chilled 1.5mL tubes and used as whole cell extracts (WCE). WCE were completed with 3.6µL 0.55M Sodium Dodecyl Sulfate (6X) and heated at 96°C for 10min. Samples were separated using a 3% acrylamide/bis-acrylamide (37.5:1) stacking gel and a 10% acrylamide/bis-acrylamide (37.5:1, Bioshop, cat#ACR005.500) resolution gel. The resolution part of the gel was transferred to a nitrocellulose membrane (BioRad, cat#162-0115) using a transfer buffer composed of 1.92M Glycine (Bioshop, cat#GLN001.5) and 0.25M Tris (Bioshop, cat#TRS001.5). The nitrocellulose membrane was blocked, in PBS supplemented with 0.5% Tween 20 (MP Biomedicals, cat#0210316890; PBS-T) containing 5% milk. Polyclonal anti-SARS-Related Coronavirus 2 Spike Glycoprotein (BEI resources, cat#NR-52947) and monoclonal anti-β-Actin (Millipore Sigma, cat#A5441) antibodies were diluted 1/10 000 and 1/6000, respectively, in PBS-T supplemented with 5% Bovine Serum Albumin (Millipore Sigma, cat# A7906). SARS Nucleocapsid protein antibody (Novus Biologicals, cat#NB100-56576) was diluted 1/2000 in PBS-T to which 5% milk was added. Membranes were washed in PBS-T after overnight incubation with the primary antibodies prior to incubation with horseradish peroxidase (HRP)-conjugated secondary antibodies (Seracare or Jackson Immunoresearch Laboratories). The Western Lightning Chemiluminescence Reagent Plus (Perkin-Elmer Life Sciences, cat# NEL104001EA) was used and detected using a LAS4000mini CCD camera apparatus (GE healthcare).

**Supplemental Table 1**

**Clinical data of the patient in the room where air samples tested for infectious SARS-CoV-2 were collected**

| Sex | Female |
| --- | --- |
| Age -years | 40-49 |
| Co-morbidity | Diabetes |
| Immunization against COVID-19 | No |
| Duration of symptoms prior to sampling - days | 7 |
| Duration of hospitalization prior to sampling - days | 3 |
| Clinical manifestations at time of sampling | - Dyspnea - Severe cough |
| Therapies at time of sampling | - Intermittent administration of oxygen by nasal canula at a rate of 2L/min - Oral codeine - Dexamethasone: 6mg orally daily – started 24h before sampling |

**Supplemental Table 2**

**Rt-qPCR analysis of SARS-CoV-2 N in the supernatant of cell culture**

| **Sample ID** | **Ct *N*** | |
| --- | --- | --- |
|  | **D0** | **D3(2)** |
| MOCK | 36.12 | 36.39 |
| SARS-CoV-2 | 27.07 | 10.74 |
| #1 | 30.58 | 33.21 |
| #2 | 32.9 | 39.24 |
| #3 | 34.53 | 36.77 |
| #4 | 33.42 | 12.39 |

**Supplemental Figure 1**

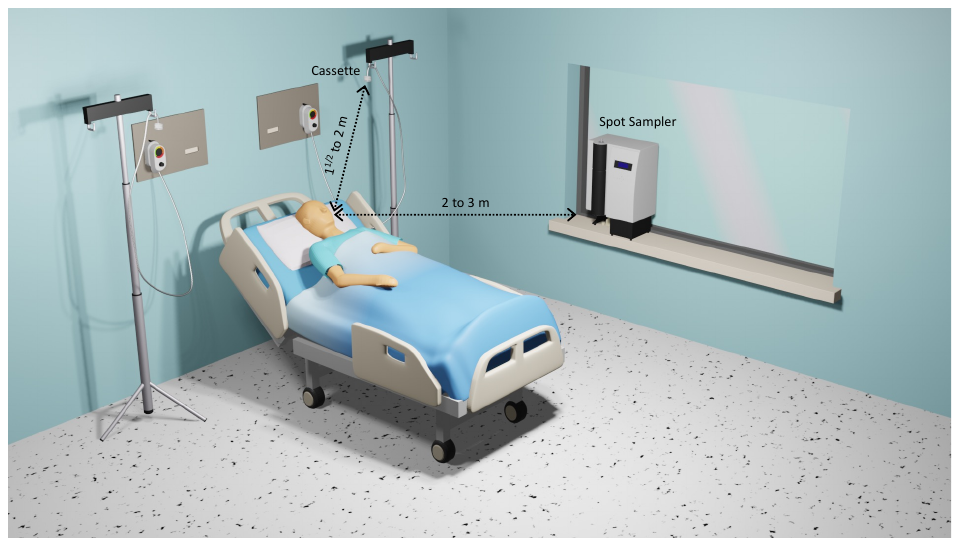
 **Supplemental Figure 1.** Hospital room of patient with COVID-19 layout depicting the position of the sampling devices and the distance to the head of the patient.
